# The sexual and gender-diverse face more health challenges during COVID-19: A large-scale social media analysis with natural language processing

**DOI:** 10.1101/2024.06.14.24308944

**Authors:** Zhiyun Zhang, Yining Hua, Peilin Zhou, Shixu Lin, Minghui Li, Yujie Zhang, Li Zhou, Yanhui Liao, Jie Yang

## Abstract

**Background:** The COVID-19 pandemic has caused a disproportionate impact on the sex and gender diversity (SGD) community. Compared with non-SGD populations, their social relations and health status are more vulnerable, whereas public health data regarding SGD is scarce. **Methods:** To analyze the concerns and health status of SGD individuals, this cohort study leveraged 471,371,477 tweets from 251,455 SGD and 22,644,411 non-SGD users, spanning from February 1, 2020, to April 30, 2022. The outcome measures comprised the distribution and dynamics of COVID-related topics, attitudes towards vaccines and the prevalence of symptoms. **Results:** Topic analysis revealed that SGD users engaged more frequently in discussions related to “friends and family” (20.5% vs 13.1%, P<0.001) and “wear masks” (10.1% vs 8.3%, P<0.001) compared to non-SGD users. Additionally, SGD users exhibited a significantly higher proportion of positive sentiment in tweets about vaccines, including Moderna, Pfizer, AstraZeneca, and Johnson & Johnson. Among 102,464 users who self-reported COVID-19 diagnoses, SGD users disclosed significantly higher frequencies of mentioning 61 out of 69 COVID-related symptoms to non-SGD users, encompassing both physical and mental health challenges. **Conclusion:** The results provide insights to an understanding of the unique needs and experiences of the SGD community during the pandemic, emphasizing the value of social media data in epidemiological and public health research.

## 1. Introduction

The COVID-19 pandemic has posed tremendous pressure on global health systems, leading to issues such as resource constraints and overcrowding in medical facilities [1]. These challenges are particularly acute for vulnerable communities, including sexual and gender diverse (SGD) individuals, who frequently face systemic inequality [2]. Prior studies have reported that individuals from these communities encounter increased barriers to healthcare access due to both overt and systemic discrimination, as well as inadequate health insurance coverage [1, 3]. They often have a higher prevalence of chronic conditions that are associated with severe COVID-19 outcomes, such as diabetes [4], cardiovascular diseases, and respiratory conditions like asthma [5, 6]. In addition, systemic factors also placed these individuals in socially or emotionally challenging environments, heightening their risk for severe mental health issues [7].

This backdrop of heightened vulnerability underscores the crucial role that vaccine availability and acceptance play in curbing the spread of COVID-19 among SGD populations [8, 9]. Existing research on vaccine hesitancy often overlooks or misrepresents these individuals [10], despite the fact that clinical and social factors contribute to their vaccine acceptance [11]. For instance, concerns over underlying health conditions [10], and considerations of vaccine efficacy and safety [12] all shape their attitudes towards vaccination. It is crucial, therefore, to target SGD individuals specifically, aiming to enhance their vaccine acceptance by deeply understanding their stance.

However, research on the health of SGD individuals during the COVID-19 pandemic faces a notable gap in both depth and breadth. Most studies rely on online surveys and questionnaires [13, 14], constrained by the inherent biases of questionnaire design and the prolonged intervals of data collection. Moreover, research utilizing electronic health records typically focuses on specific symptoms of severe COVID-19, influenced by hospital admission rates and delays in gathering SGD data [15, 16], thus lacking in generalizability. Additionally, the access to electronic health records is highly restricted, limiting the coverage of patients [17]. A major challenge in conducting comprehensive health status analysis for SGD populations lies in selecting large-scale, representative cohorts, which underscores the potential of alternative data sources, such as social media, which has been increasingly validated as a valuable tool in public health research [18–20]. Social media-related studies cover topics ranging from mental healthcare [21–23] to disease symptoms [24] and public acceptance of treatments [25–27], both in the context of COVID-19 and previous health crises such as H1N1 [28] and Zika [29]. Meanwhile, new deep learning-based language models, pipelines, and datasets [30–33] offer opportunities to analyze the massive textual information from social media platforms. This synergy between natural language processing (NLP) and social media analytics opens up novel avenues for research that span both data collection and analytical interpretation [18].

Within this context, our study leverages large-scale Twitter data and NLP methodologies to scrutinize the health and well-being of SGD individuals during the COVID-19 pandemic. We address three principal research questions: 1) What are the predominant topics discussed by SGD Twitter users during the pandemic? 2) How concerned are SGD individuals about pandemic precautions, such as mask-wearing and vaccination? 3) Do SGD individuals face more acute symptom risks and mental health challenges compared to non-SGD individuals during the pandemic?

To address the questions above, we employ Latent Dirichlet Allocation (LDA) models to discern public discussion themes and track their temporal evolution. Named Entity Recognition (NER) and Targeted Sentiment Analysis (TSA) models—both grounded in advanced NLP techniques and trained on Twitter-specific datasets—are used to compare vaccine perceptions between SGD and non-SGD individuals. We also identify and analyze Twitter users who have self-reported a COVID-19 diagnosis to compare health outcomes across SGD and non-SGD groups. Our preliminary results underline that SGD individuals manifest significantly elevated symptomatology and mental health challenges, emphasizing an imperative for specialized interventions.

## 2. Methods

### Experimental Design

This cohort study collected a comprehensive dataset from February 2020 to April 2022, with the data collection process adhering to Twitter’s Terms of Service. Ethical approval was secured from the Institutional Review Board of Zhejiang University and Mass General Brigham. An overview of data distribution and study design is provided in **Figure S1**. SGD users were identified through user profiles, and topic modeling techniques were employed to analyze the content. Further statistical analyses were performed to understand their sentiments regarding COVID-19 vaccines, compare self-reported symptoms between SGD and non-SGD users, and investigate their mental health status.

### Data collection and selection

This study collected tweets through leveraging tweet IDs from a public coronavirus Twitter dataset [34], which follows specified accounts and collects real-time tweets mentioning specific keywords. We instituted a filtering process where tweets containing URLs were excluded to attenuate the impact of news and automated bot activities. Subsequently, we focused on identifying tweets from SGD users, including lesbian, gay, bisexual, transgender, queer, intersex, and asexual individuals [35]. SGD users were filtered through user profiles using keyword filtering and regular expression matching (**Table S1**): 1) User profiles must contain SGD-related keywords. 2) There should be no negation words before or after the keywords. 3) The keywords should not be preceded or followed by terms such as “advocator” and “supporter” as some users may advocate for SGD rights without necessarily being SGD themselves. A manual validation process was conducted on a subset of 500 selected SGD users, achieving a classification accuracy of 93.8%. We also evaluated the baseline characteristics of geographic information on the validation subset (**Table S2**).

### Statistical Analysis

All statistical tests were conducted using Python 3.8, and were two-tailed tests, with significance levels adjusted using Bonferroni corrections for multiple comparisons.

#### Topic modeling

Given the imbalanced dataset with a disproportionate number of tweets from non-SGD users, we performed a random under-sampling to achieve parity in tweet numbers (n=2,296,289 for both groups) and sensitivity analysis was applied to verify the stability of the under-sampling (Supplementary methods and **Table S4**). The random sampled tweets were then preprocessed through: 1) removing the mention symbol “@” and the quoted usernames, 2) removing stop and short words with less than two letters, 3) applying word lemmatization, 4) adding bigrams and trigrams that co-occur more than five times, and 5) removing short tweets containing less than five tokens.

After preprocessing, 3,498,468 tweets were subjected to Latent Dirichlet Allocation (LDA) [36] using the Gensim package [37]. Model selection criteria included both topic coherence and model perplexity, tested over a range of 10 to 50 topics. The topic number was set to 12 in our case according to experiments on balancing coherence and perplexity scores (**Figure S2**). Topic validity was further confirmed through visualization using pyLDAvis [38] and manual inspection of the top 20 keywords per topic (**Table S3**). To compare the discussion differences between SGD users and non-SGD users on a specific topic, we applied *Scattertext* [39] to visualize the word frequency.

#### Sentiment Analysis of vaccines

We used a pre-trained language model, COVID-Twitter-BERT [30], which was a BERT-LARGE structure pre-trained on 160 million COVID-19-related tweets, as the backbone for our named entity analysis (NER) and target sentiment analysis (TSA) models. A linear layer and a SoftMax function were added to the end of CT-BERT [40] to predict the span of each vaccine entity for the NER model. The encoder of BERT-SPC [41] was replaced with CT-BERT for the TSA model. The models were fine-tuned on the training set of the Medical Entities and Targeted Sentiments on COVID-19-related tweets (METS-CoV) dataset [32] using NCRF++ [42], both are part of our prior work. This dataset included annotations for vaccine entities and their corresponding sentiment labels in tweets. The performance of the NER and TSA models was tested on the vaccine entity from the METS-CoV test set and resulted in an F1 score of 90.44% and an accuracy of 79.15%, respectively. As most of the recognized vaccine entities were informal expressions or misspelled, we manually incorporated the expressions of vaccine entities (details provided in **Table S5**) and selected the four most frequently mentioned COVID-19 vaccines (Moderna, Pfizer, AstraZeneca, and Johnson & Johnson) for in-depth analysis.

#### Symptom extraction and identification

Tweets that self-reported COVID-19 diagnoses were identified using lexicon filtering (details provided in **Table S6**). Tweets not written in the first person or contained negative or uncertain expressions (e.g., wonder, thought, might, etc.) before and after the keywords were filtered out through regular matching. For each selected user, we determined the date of diagnosis based on the content of the first self-report tweet. If the date of diagnosis was not specified in that tweet, we assumed that the time of tweeting was the time of diagnosis.

Tweets posted before and after 30 days of their self-report date were collected for users who self-reported COVID-19 diagnoses. We then screened these tweets using a COVID-19 symptom lexicon developed by Wu et al. [43], which contains commonly used synonyms and colloquial variants on social media that pertain to symptoms and their associated affected organs or systems. For the identification of mental health-related tweets, we utilized an exhaustive mental health lexicon [44] which has been rigorously validated by professionals in the fields of psychiatry and psychology. This lexicon encompasses 231 keywords distributed across four major mental health conditions: anxiety, depression, insomnia, and substance use disorders.

## 3. Results

We downloaded a total of 471,371,477 tweets from the public COVID-19 Twitter dataset. After excluding tweets with URLs, the dataset was narrowed down to 169,669,346 tweets. Within this set, 2,296,289 tweets originated from 251,455 SGD users and 167,373,057 tweets originated from 22,644,411 non-SGD users (details provided in **Table S1**).

### Topic distribution and discrepancies

After preprocessing, 3,498,468 tweets were subjected to the topic model, and the top 5 most discussed topics were “friends and family” (16.8%, 95% CI - 16.7% to 16.9%), “lockdown” (11.4%, 95% CI – 11.3% to 11.5%), “vaccine” (10.7%, 95% CI – 10.6% to 10.8%), “politics” (10.7%, 95% CI – 10.6% to 10.8%), and “wearing masks” (9.2%, 95% CI – 9.1% to 9.3%). Notably, these trends varied over time and exhibited specific peaks, as smoothed by a 7-day moving average (**Figure 1A**). Besides, topic fluctuations reflect significant events during the pandemic. For instance, the sharp increase in discussions on “gender and race” in May 2020 corresponds to heightened concerns about racial equity [45] following the police killing of George Floyd, an unarmed Black civilian [46]. Similarly, the U.S. presidential election and subsequent debates over COVID-19 policies [47] caused a spike in the topic “politics” in November 2020.

**Fig. 1.**
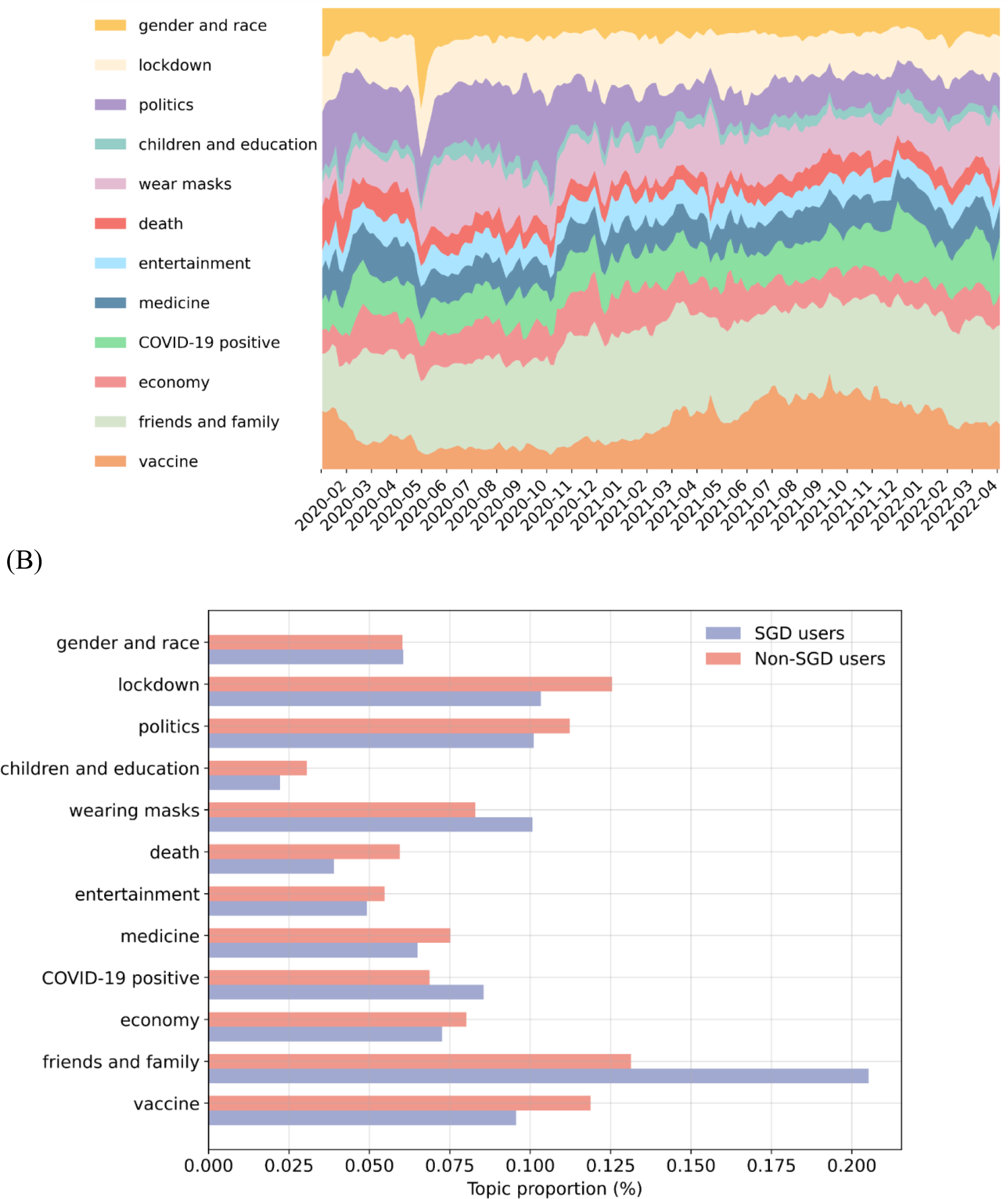
Topic distributions in COVID-19-related tweets. (A) The topic proportion distribution of SGD user-posted tweets over time. (B) Topic distributions of tweets posted by SGD users and non-SGD users.

When comparing topic frequencies between SGD and non-SGD users using χ² test with Bonferroni adjusted significance level P < 0.004, the data indicate that discussions about “friends and family” were significantly more prevalent among the former (20.5% vs 13.1%, P<0.001) (**Figure 1B**). For a deeper insight into the discussion variations, we used *Scattertext* [39] to visualize the word frequencies (**Figure S3**). The result highlights that terms regarding family members occur frequently, with SGD users often mentioning “partner” while non-SGD users more frequently use terms like “daughter” and “baby”. Furthermore, SGD users express a range of emotions more frequently, especially negative ones like “anxiety”, “upset”, “angry”, “depression”. For other topics, SGD users were more likely to talk about “wear masks” (10.1% vs 8.3%, P<0.001) and “COVID-19 positive” (8.6% vs 6.9%, P<0.001), while non-SGD users discussed other topics like “vaccine” (9.6% vs 11.9%, P<0.001) and “lockdown” (10.3% vs 12.5%, P<0.001) more often.

### Attitude Towards COVID-19 Vaccines

**Table 1** shows the distribution of three-category sentiments (positive, neutral, and negative), wherein the majority were characterized as neutral. Notably, sentiments toward the Pfizer vaccine exhibited the highest frequency of positive evaluations. Contrarily, attitudes toward the AstraZeneca vaccine appeared the most polarized among SGD users when contrasted with non-SGD users. Utilizing an Independent Samples t-test for statistical analysis, we found that SGD users displayed significantly higher proportions of positive sentiments for all four vaccine types including Moderna (tweets no. [%]. 610 [12.6] vs 28828 [8.6], P<0.01), Pfizer (tweets no. [%]. 984 [12.9] vs 58109 [8.8], P<0.01), AstraZeneca (tweets no. [%]. 158 [11.0] vs 16227 [3.8], P<0.01), and Johnson & Johnson (tweets no. [%]. 208 [9.3] vs 11482 [6.5], P<0.01). Furthermore, the proportions of negative sentiments for both the Moderna (tweets no. [%]. 182 [3.7] vs 21271 [6.4], P<0.01) and Pfizer (tweets no. [%]. 327 [4.3] vs 51669 [7.8], P<0.01) vaccines were significantly lower among SGD users compared to the non-SGD group.

**Table 1.** Percentage of positive and negative sentiments in vaccine-related tweets.

| Vaccine | Positive |  |  | Negative |  |  |
| --- | --- | --- | --- | --- | --- | --- |
|  | Tweets, No. (%) [95% CI<sup>a</sup>] |  | P value<sup>b</sup><br>(SGD greater) | Tweets, No. (%) [95% CI] |  | P value<br>(SGD smaller) |
|  | SGD | Non-SGD |  | SGD | Non-SGD |  |
| **Moderna** | 610 (12.6)<br>[11.6-13.5] | 28828 (8.6)<br>[8.5-8.7] | <0.001 | 182 (3.7)<br>[3.2-4.3] | 21271 (6.4)<br>[6.3-6.5] | <0.001 |
| **Pfizer** | 984 (12.9)<br>[12.2-13.7] | 58109 (8.8)<br>[8.7-8.8] | <0.001 | 327 (4.3)<br>[3.8-4.8] | 51669 (7.8)<br>[7.7-7.9] | <0.001 |
| **AstraZeneca** | 158 (11.0)<br>[9.4-12.7] | 16227 (3.8)<br>[3.8-3.9] | <0.001 | 68 (4.8)<br>[3.7-5.9] | 11071 (2.6)<br>[2.6-2.7] | 0.999 |
| **Johnson & Johnson** | 208 (9.3)<br>[8.1-10.5] | 11482 (6.5)<br>[6.4-6.6] | <0.001 | 127 (5.6)<br>[4.7-6.6] | 11191 (6.3)<br>[6.2-6.4] | 0.101 |

<sup>a</sup> We used the ratio t-test to calculate 95% confidence intervals and the independent samples t-test to calculate significance.
<sup>b</sup> Significance were set at $P < .006$ after Bonferroni correction.

### Physical and mental health status

We identified 2,098 SGD and 100,366 non-SGD users who self-reported COVID-19 diagnoses (the overview of users’ filtering process is provided in **Figure S4**). Analysis of tweets within a 30-day window surrounding the self-reported date yielded mentions of 69 unique symptoms, implicating 15 distinct organ systems or physiological functions (**Supplementary File 1**). An independent samples t-test showed that the frequency of mentions for 61 of these 69 symptoms was significantly higher (P < 7.25×10^-4^, Bonferroni adjusted) among the SGD cohort compared to the non-SGD group.

We then calculated mention rates for each symptom in both groups. **Figure 2A** displays mention rates for symptoms cited by more than 1,000 individuals in the SGD group and 35,000 in the non-SGD group. Symptoms most frequently mentioned—such as anxiety, nausea, and allergic reactions—had higher prevalence among SGD individuals. **Figure 2B** further shows that mention rates for symptoms related to mental and musculoskeletal health were especially elevated in the SGD individuals, followed by mental symptoms, trachea and lung, and brain.

**Fig. 2.**
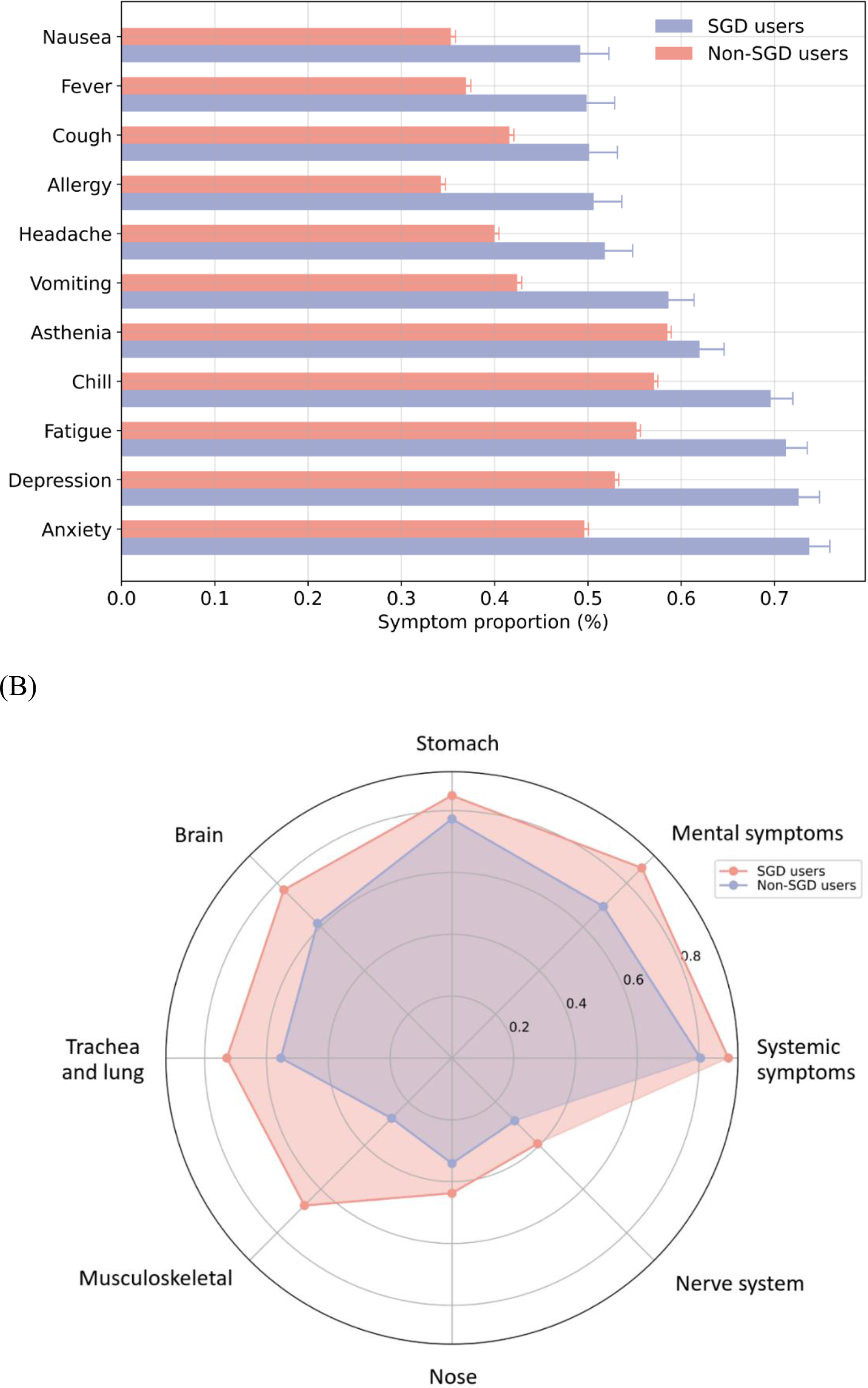
Symptom distributions in users who self-report COVID-19 positive. (A) Symptom distributions of SGD and non-SGD Twitter users. (B) Distribution of the eight most affected organs or systems of SGD and non-SGD Twitter users.

For mental health analysis, from 137,860 mental health-related tweets contributed by SGD individuals and 7,647,024 tweets contributed by non-SGD individuals, we identified 1,984,317 tweets related to anxiety, 5,258,324 related to depression, 586,648 related to insomnia, and 200,905 related to substance use disorders.

Figure 3 shows the temporal distribution of these mental health-related tweets. We observed an initial surge in tweets concerning anxiety, depression, and substance use at the onset of the pandemic, followed by a stabilization to baseline levels. Contrastingly, the proportion of tweets pertaining to insomnia demonstrated a continual increase over time. Given that the tweet distribution over time deviated from normality, we employed two-tailed Wilcoxon matched-pairs signed-ranks tests (**Table 2**), and it turned out that SGD users exhibited higher prevalence of anxiety (% tweets median [IQR]. 1.58 [0.53] vs 1.05 [0.16], P<0.01), depression (% tweets median [IQR]. 3.63 [0.89] vs 3.02 [0.31], P<0.01), insomnia (% tweets median [IQR]. 0.52 [0.33] vs 0.32 [0.15], P<0.01), and addiction (% tweets median [IQR]. 0.13 [0.10] vs 0.12 [0.02], P<0.01) symptoms.

**Fig. 3.**
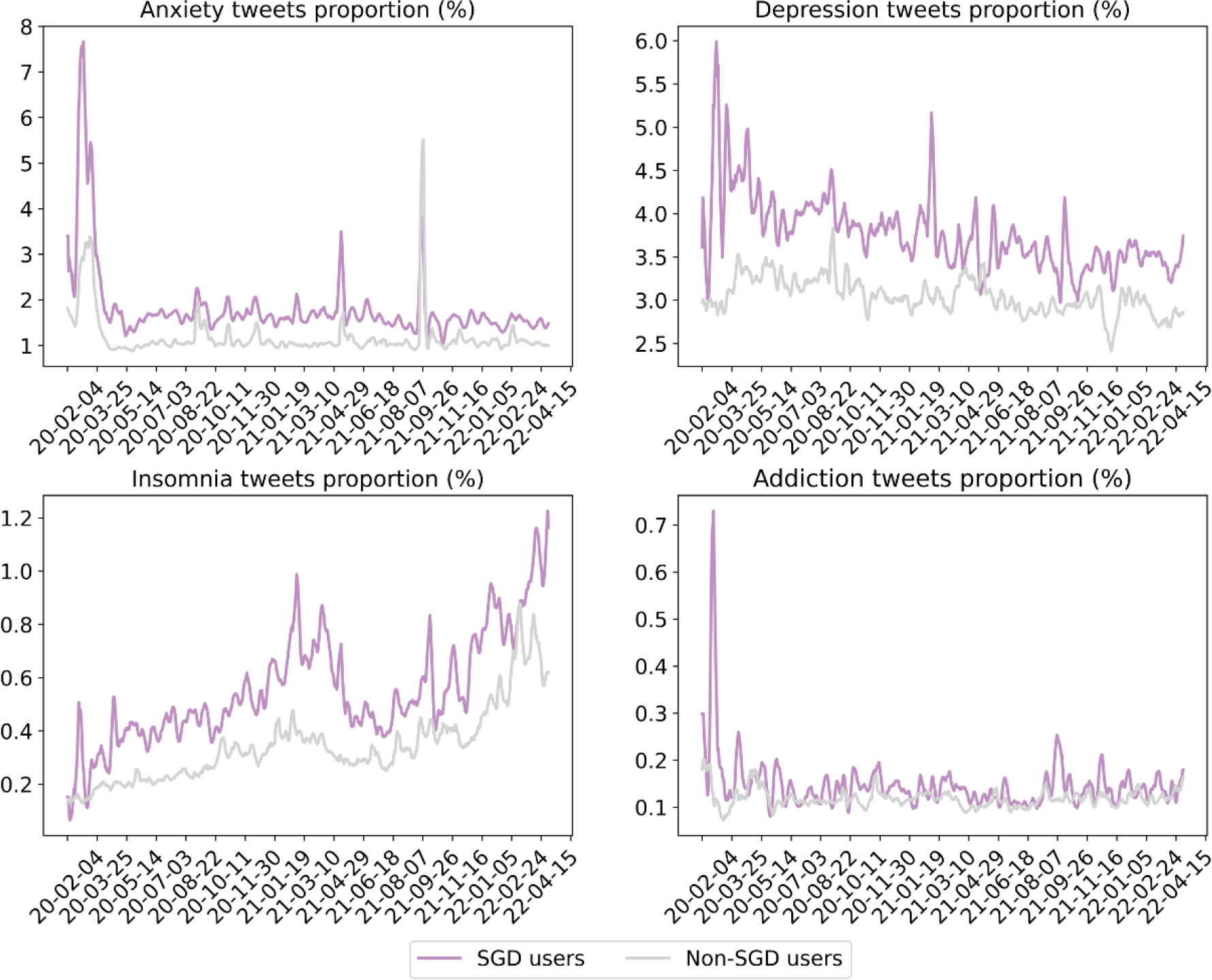
The proportion of tweets concerning mental health to the total number of tweets. We used a 7-day moving average to smooth the curve.

**Table 2.** The daily proportion of mental health-related tweets of SGD and non-SGD users. W represents the sum of the ranks of the differences above zero.

| Mental symptoms | SGD (% tweets), median(IQR<sup>a</sup>) | Non-SGD (% tweets), median (IQR) | W | P-value<sup>b</sup> |
| --- | --- | --- | --- | --- |
| anxiety | 1.585 (0.530) | 1.047 (0.158) | 317574 | <0.001 |
| depression | 3.634 (0.885) | 3.024 (0.315) | 302044 | <0.001 |
| **insomnia** | 0.523 (0.325) | 0.320 (0.154) | 312525 | <0.001 |
| **addiction** | 0.127 (0.103) | 0.115 (0.024) | 203679 | <0.001 |

<sup>a</sup> We used the IQR and Wilcoxon matched-pairs signed-ranks test in the statistical analysis.
<sup>b</sup> Significance were set at $P < .016$ after Bonferroni correction.

## 4. Discussion

In this pioneering social media-based retrospective cohort study, we examined the differential impact of the COVID-19 pandemic on SGD individuals by analyzing a large corpus of pandemic-related tweets over two and a half years. Our methodological approach encompasses: 1) the use of topic modeling to delineate concerns unique to SGD individuals; 2) the application of advanced deep learning-based NLP algorithms for sentiment analysis towards vaccines; and 3) a comparison of self-reported COVID-19 symptoms between SGD and non-SGD individuals. Through these avenues, our research aims to elucidate the unique challenges confronting SGD individuals during the pandemic and to inform targeted interventions designed to alleviate their physical and psychological burdens, and therefore enhance their well-being.

Our topic modelling results divulge a heightened focus among SGD individuals on themes like “friends and family”, where they tend to express negative emotions more frequently. Research indicates that pandemic policies, such as school closures and lockdowns, have severed some social connections, leaving SGD individuals more dependent on family members [3]. However, older individuals are often isolated as they are four times less likely to have children and SGD youth are forced to be at home with unsupportive parents [7]. Emotional and mental health harms may arise from the lack of supportive surroundings. We also observed that SGD individuals are more likely to discuss topics related to preventative health measures such as “mask-wearing” and “COVID-19 testing.” These observations align with previous research by Sears Brad et al. conducted in the US [48]. They found that SGD individuals prefer to wear masks (94.0% vs. 89.9%) and take COVID-19 tests (38.3% vs. 29.0%).

Our sentiment analysis further shows more positive attitudes towards vaccines within SGD individuals compared to their non-SGD counterparts, which is positively correlated with stronger vaccination stance [49, 50]. This higher rate of vaccine adoption is corroborated by telephonic surveys and suggests greater compliance with public health directives within these communities [51, 52]. These results suggest that SGD individuals exhibit a greater awareness of the importance of precautionary measures and prefer to comply with public health orders during the pandemic [53]. The higher willingness of SGD individuals to vaccinate may be linked to their altruistic tendencies [54] and higher levels of perceived health vulnerability [55]. Nonetheless, vaccination rates for SGD populations vary widely over different regions and ethnicities [10]. More efforts are needed to assess vaccination rates in these areas and improve the coverage for SGD individuals without insurance or documents.

In examining self-reported symptoms among COVID-19-positive individuals, our data reveals concordance with clinical studies [56–58] regarding the most frequently mentioned symptoms. However, we found a higher frequency of certain mild symptoms such as musculoskeletal and mental health issues within the SGD population, as compared to electronic health record-based studies [59]. This underscores the utility of social media as a complementary data source for capturing a broader spectrum of patient experiences that might not be adequately recorded in clinical settings. In addition, we noted that SGD individuals are more likely to experience more severe symptoms after COVID-19 infection. This may be due to inadequate health insurance coverage and higher-than-average rates of underlying diseases such as diabetes, and asthma, which can increase the risk of severe symptoms [5, 6]. Besides, SGD users mentioned musculoskeletal symptoms (body pain, myalgia pain, arthralgia pain, etc.) at a particularly higher rate compared to non-SGD users. These symptoms are often associated with severe disease as they can be triggered by increased inflammatory factors (e.g., interleukin-6) during infection [60, 61].

The psychological ramifications within SGD communities warrant nuanced attention, as our study indicates elevated rates of mental health symptoms than non-SGD groups during the pandemic, which is consistent with pre-existing literature employing the PHQ4 scale and online surveys [13, 14, 62]. In terms of temporal variations, the frequency of insomnia-related tweets exhibited a correlation with diagnosed COVID-19 cases, peaking in January 2021 and rising steadily from January through April 2022. These trends are congruent with clinical literature suggesting a high correlation between insomnia and COVID-19 infection [63–65]. Conversely, fluctuations in tweets related to other mental health conditions— namely depression, anxiety, and addiction—appeared to be more significantly influenced by social determinants. For instance, a sudden spike in anxiety-related tweets occurred in September 2021, and the majority of the discussion was focused on the increase in fuel prices. This phenomenon has also been observed by previous research [44]. During the pandemic’s initial outbreak in February 2020, SGD communities experienced more pronounced spikes in symptoms of depression, anxiety, and addiction compared to their non-SGD counterparts. These exacerbated symptoms may be attributed to distinct and more severe social challenges confronting SGD individuals, such as limited access to supplies and healthcare [66]. Moreover, it is crucial to highlight the constrained social support networks often associated with SGD communities, which include family, partners, and peers. Such networks frequently lack the resilience and social capital to act as effective buffers against the immediate repercussions of both social changes and health crises [67].

We acknowledge several limitations in our study. First, the age and geographical distributions of Twitter users are skewed, introducing potential selection bias that may limit the external validity of our findings. For instance, individuals with lower socio-economic status or those of advanced age may be underrepresented on Twitter, thereby introducing a bias towards certain demographic groups [68]. Secondly, despite the application of advanced natural language processing models employing deep learning, our pipeline is susceptible to misclassification bias due to lexical ambiguity [69]. To assess this issue, we conducted a random selection of 500 tweets identified as originating from SGD individuals. Manual validation of these tweets suggests that 31 (6.2%) tweets may have been inaccurately categorized. Moreover, the composition of our non-SGD control group is subject to information bias; SGD users who have not publicly disclosed their identities on Twitter might be included, which could attenuate the observed effect sizes and affect the internal validity of our findings. Furthermore, our study is confined by the absence of pre-pandemic baseline data, largely due to Twitter’s data-sharing constraints. This results in a lack of temporal control, making it challenging to differentiate the health disparities between SGD and non-SGD groups directly attributable to the pandemic. Strict filtering criteria for users that self-reported COVID-19 positive may lead to a lower recall rate, resulting in selection bias among the remaining samples and an inability to represent the entire population. Moreover, the collected symptom descriptions may be subjective to the user and lack the strictness of evidence-based medicine, but they can serve as an auxiliary tool for public health analysis. These limitations should underscore the need for cautious interpretation.

## 5. Conclusion

In summary, this pioneering study employs various NLP techniques like NER, TSA and LDA models, to provide an in-depth understanding of the experiences and health outcomes of SGD individuals during the COVID-19 pandemic. Our findings emphasize the importance of enhancing social and legal support for SGD individuals and informing public health interventions to address disparities during challenging times. The methodology and pipeline developed in this study can be applied to monitor the health of other populations, providing data-driven insights for more comprehensive public health services.

## Data Availability

The codes used in this study can be accessed at https://github.com/zooay-zzy/COVID-twitter-SGD-analysis.

## Ethical Approval

IRB approved by School of Public Health, Zhejiang University ZGL202201-2

## Author contributions

Conceptualization: J.Y., Z.Z., and Y.L. Methodology: Z.Z., S.L., and Y.Z. Investigation: Z.Z., J.Y., and M.L. Writing—original draft: Z.Z. Writing— review & editing: J.Y., Y.L., P.Z., L.Z. Z.Z. takes responsibility for the integrity of the work.

## Acknowledgments

none

## Supplementary Materials

Methods

Figures S1 to S4

Tables S1 to S6

## Methods

### Sensitivity analysis of under-sampled non-SGD tweets in the topic model

To assess the impact of different random seeds on topic distribution, we created two new samples of non-SGD tweets: Sample A, with the same number as the original sample (n = 2,296,289), and Sample B, with twice that number (n = 4,592,578). After preprocessing, 1,742,444 tweets remained in Sample A and 3,484,880 in Sample B. Then we evaluated the distribution of topics in these new corpora using the trained LDA model and compared it with the old distribution using chi square test (**Table S4**).

---

**Figure S1.**
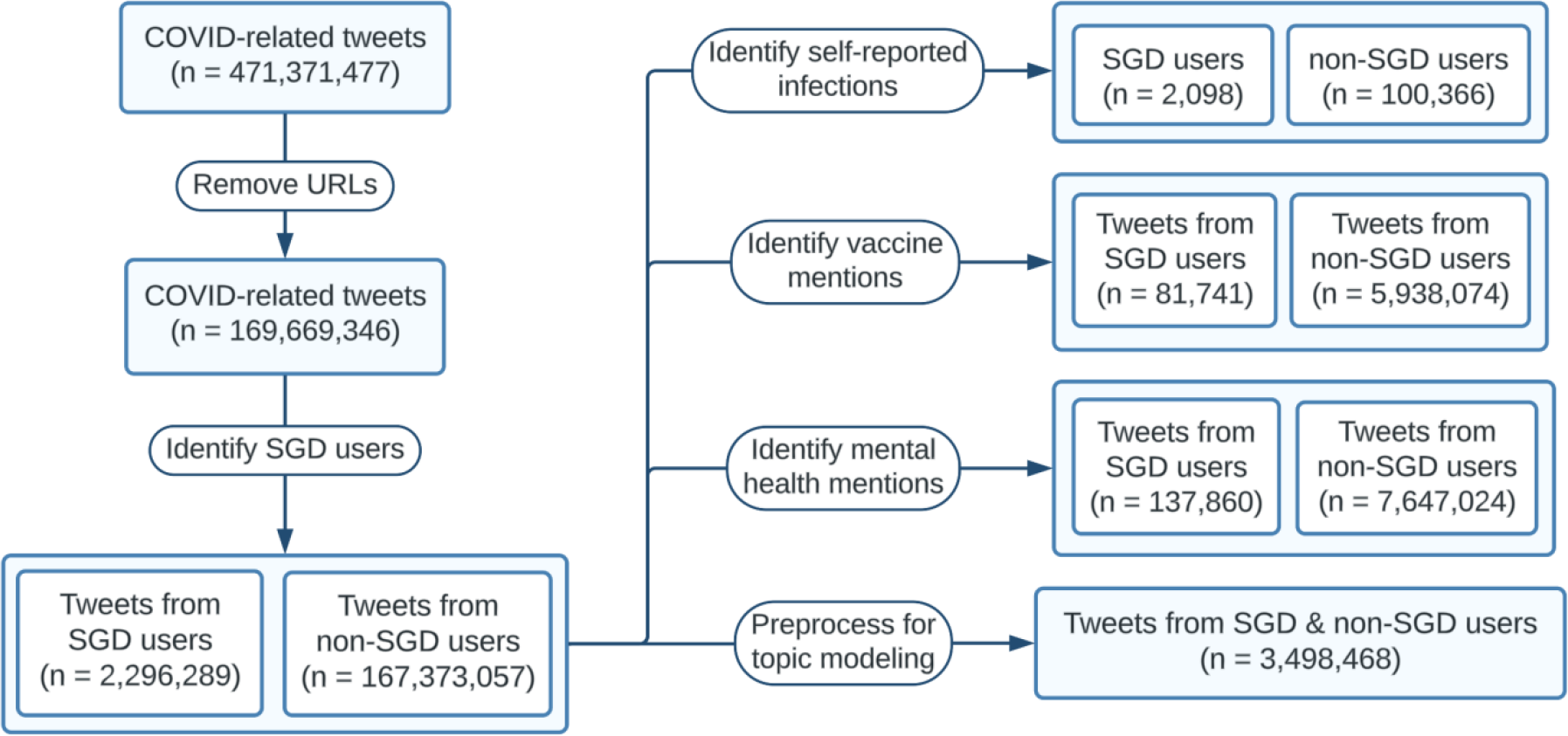
Data collection and distribution.

**Figure S2.**
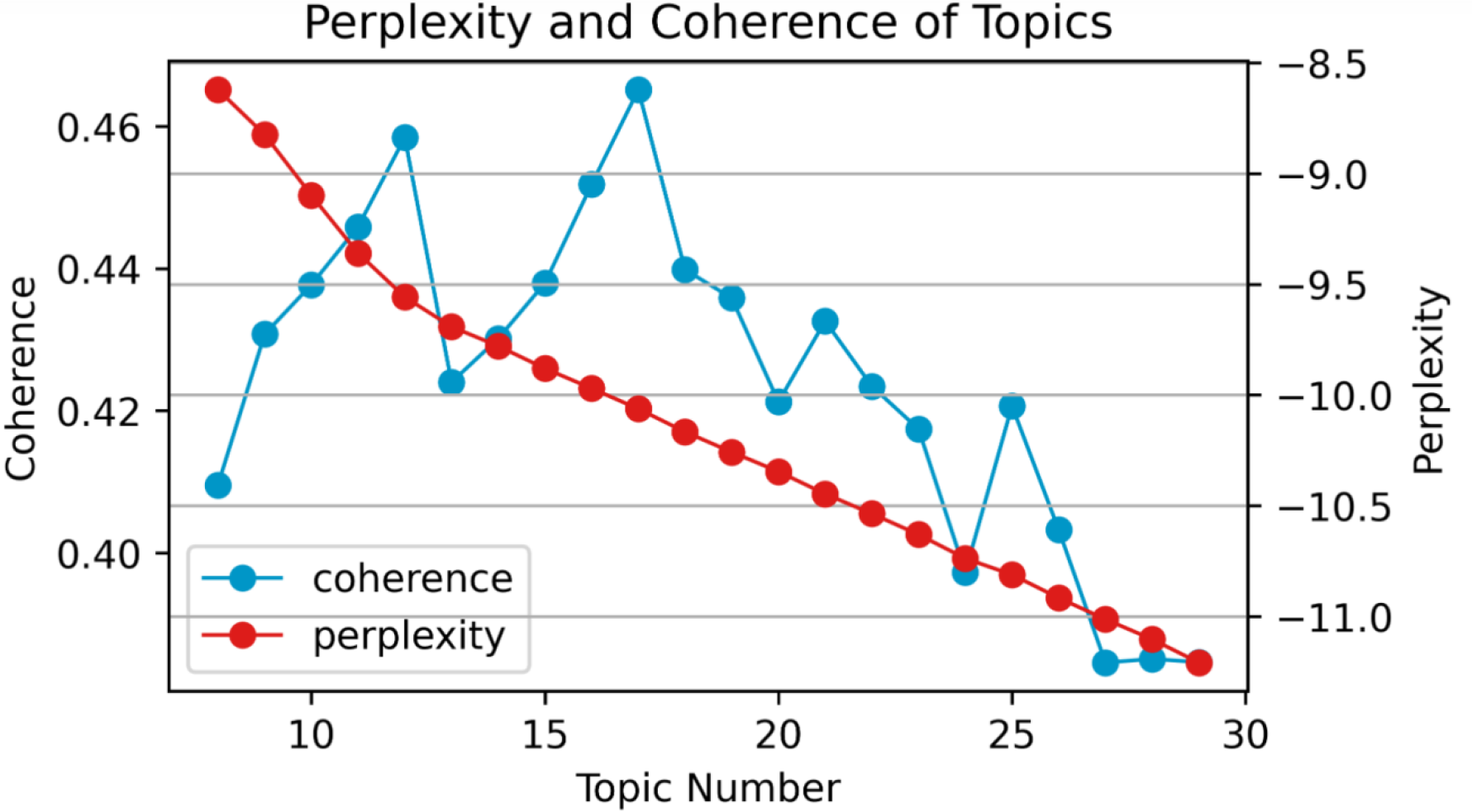
Perplexity and coherence score of different topics. Topic perplexity measures the generalizability of models to unseen data, and coherence measures the degree of semantic similarity between high-scoring words in topics.

**Figure S3.**
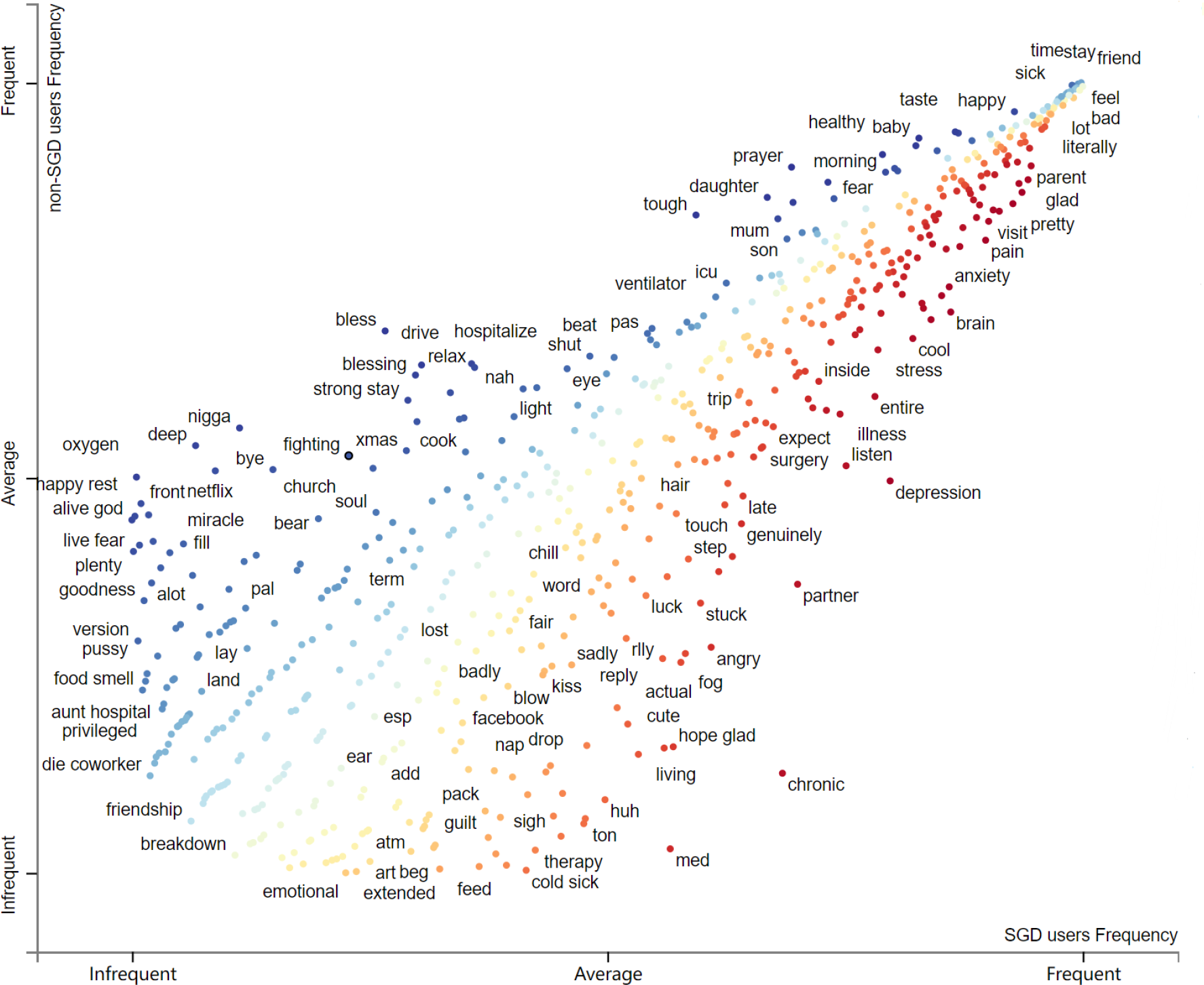
Visualization of word frequency in the topic “friend and family” using *Scattertext*. The x- and y-axes of terms are the dense ranks of their usage by SGD and non-SGD users respectively.

**Figure S4.**
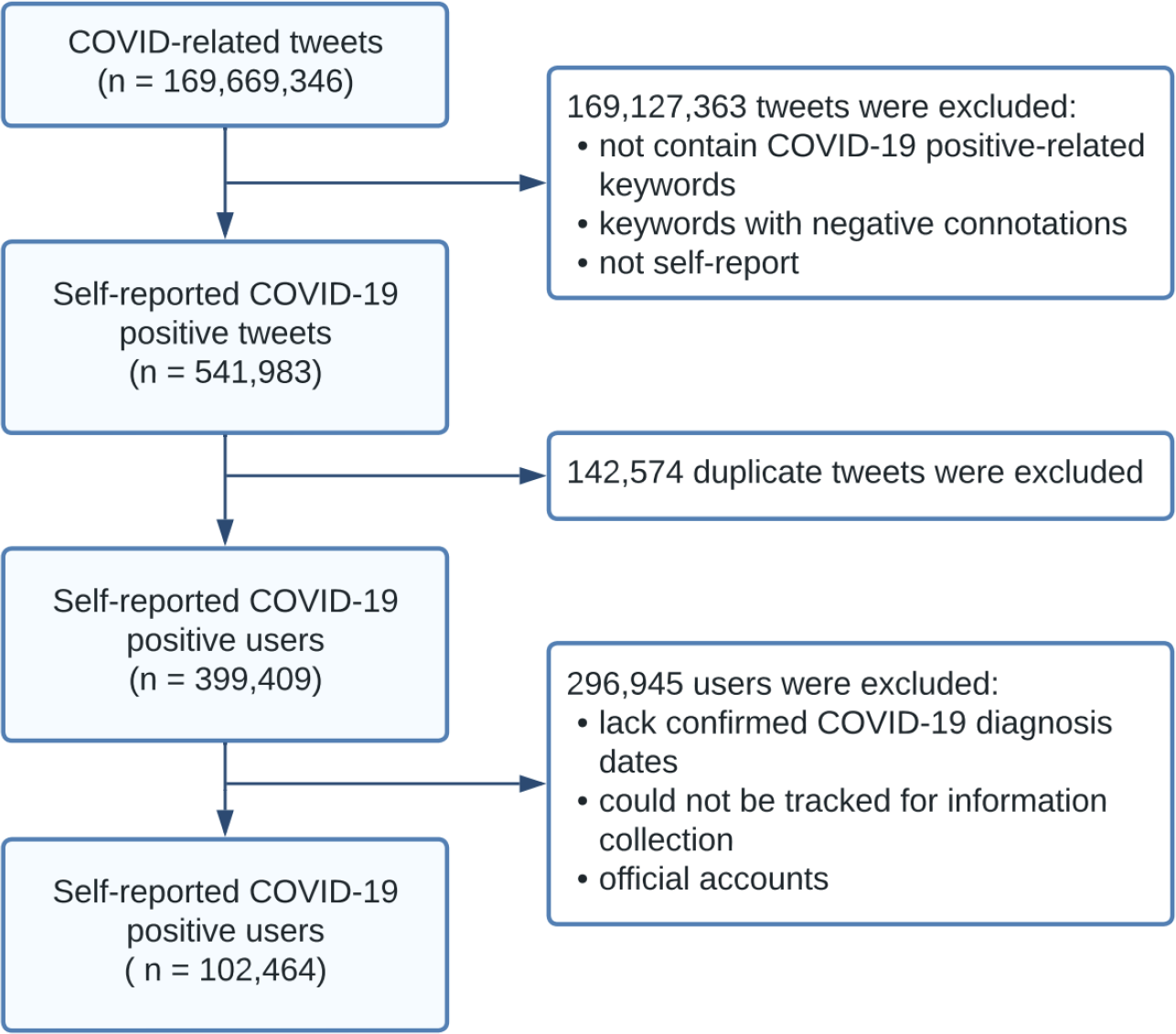
Self-reported COVID-19 positive users filtering process.

**Table S1.** Lexicon and rules for SGD selection. We used regular expressions to remove tweets with stop words before and after the SGD-related keywords to filter out negative expressions and SGD supporters but non-SGD individuals. “|” is used as a delimiter between lexicons.

| Stop words in front | SGD lexicon | Stop words behind |
| --- | --- | --- |
| not \| no \| none \| anti \| hate \| against \| pro \| advocate | lgbt \| lesbian \| gay \| bisexual \| transgender \| queer \| intersex \| asexual \| gender minority \| sex minority \| sex and gender minority | anti \| ally \| supporter \| advocator \| friendly |

**Table S2.** Geographic distribution of SGD and non-SGD users on the validation subset. The geographic information is extracted from the “place” field in tweet metadata but not the “location” field in “user” description, since the former is generated from GPS data.

|  | SGD users (%)<br>(n=500) | non-SGD users (%)<br>(n=500) | P value* |
| --- | --- | --- | --- |
| **Country** |  |  | 0.015 |
| United States | 295 (59.0) | 272 (54.4) |  |
| United Kingdom | 102 (20.4) | 98 (19.6) |  |
| Canada | 24 (4.8) | 22 (4.4) |  |
| Australia | 19 (3.8) | 18 (3.6) |  |
| India | 60 (12.0) | 90 (18.0) |  |

\*P values were calculated using $\chi^2$ test.

**Table S3.**
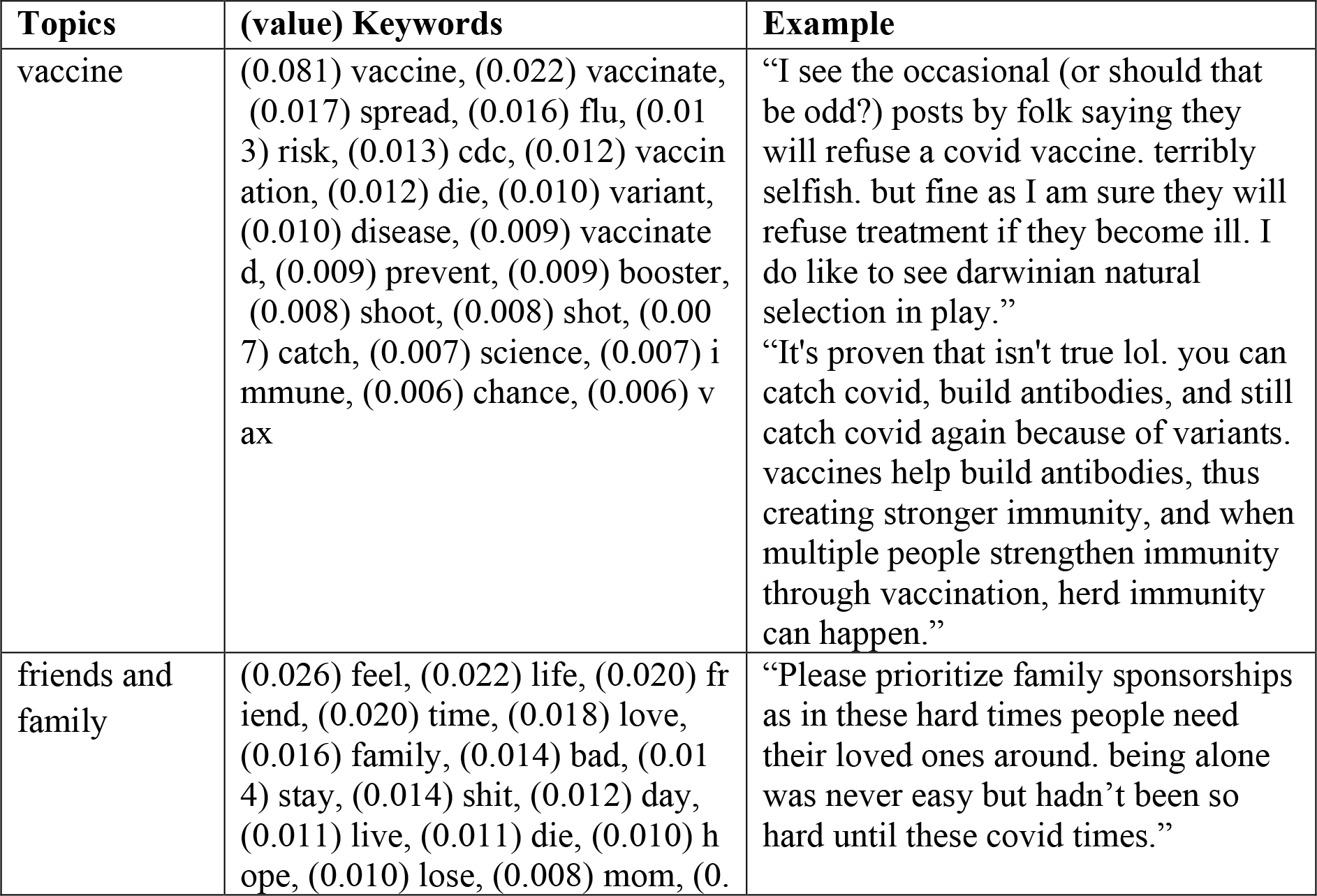
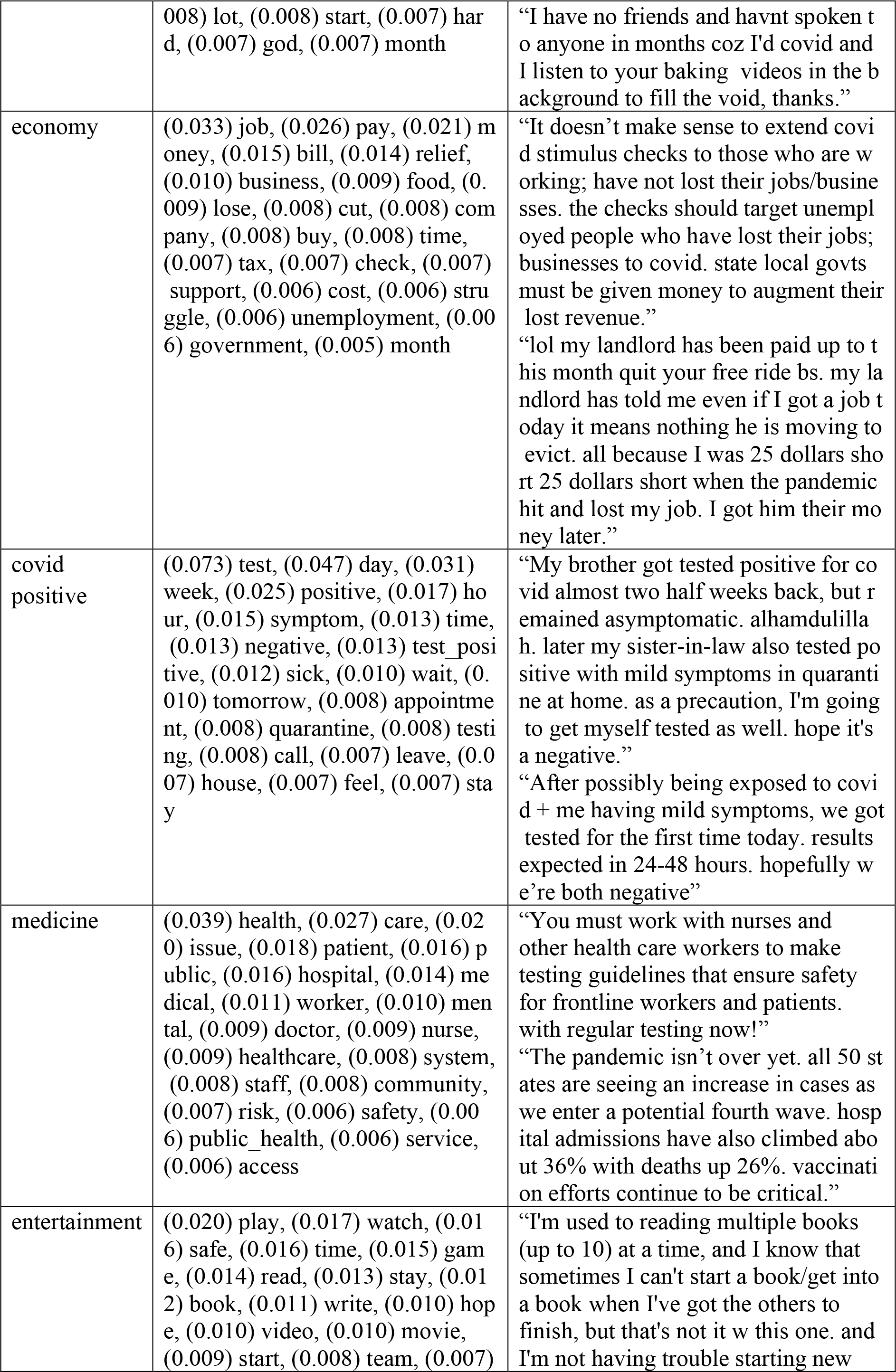
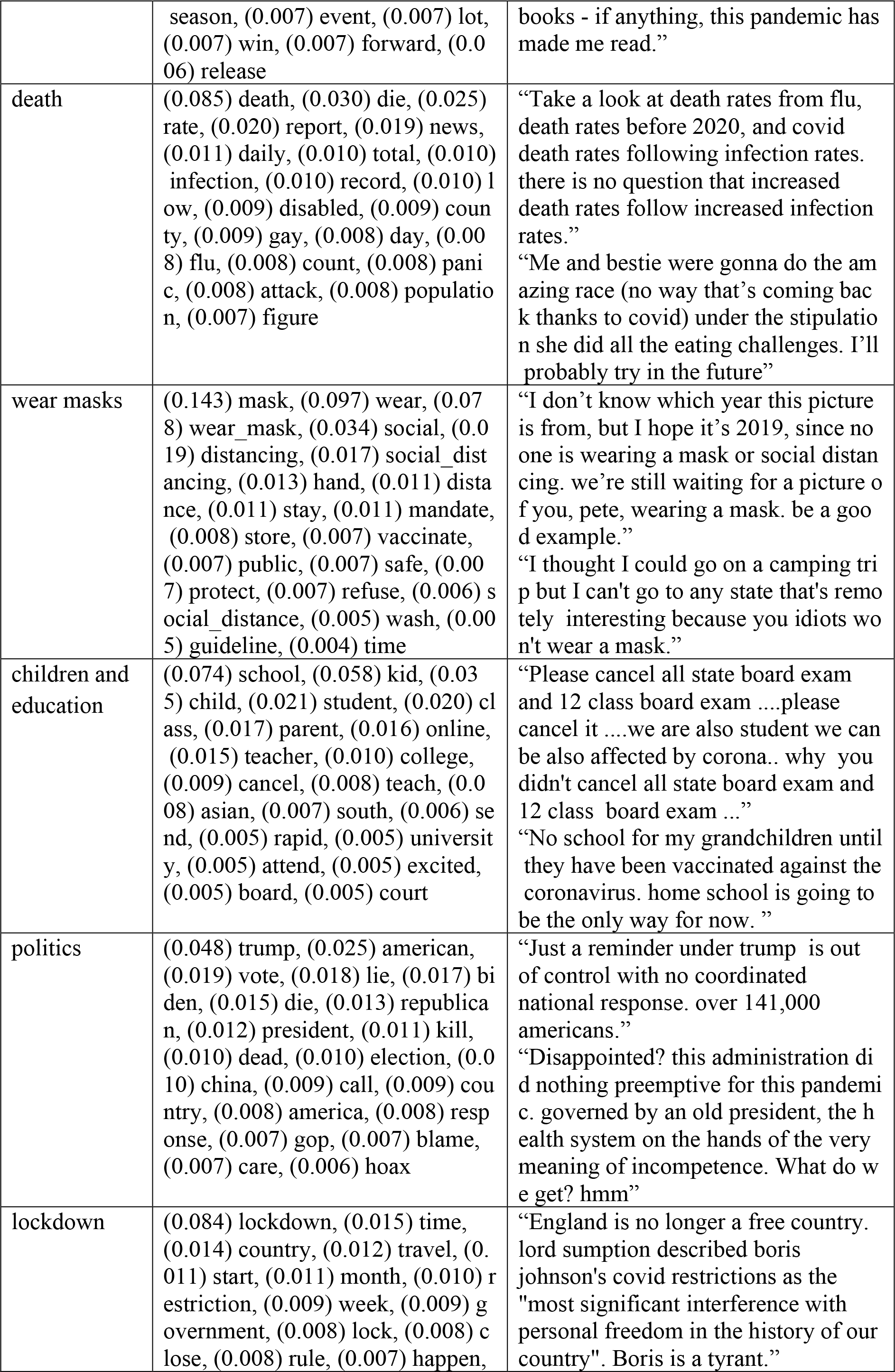
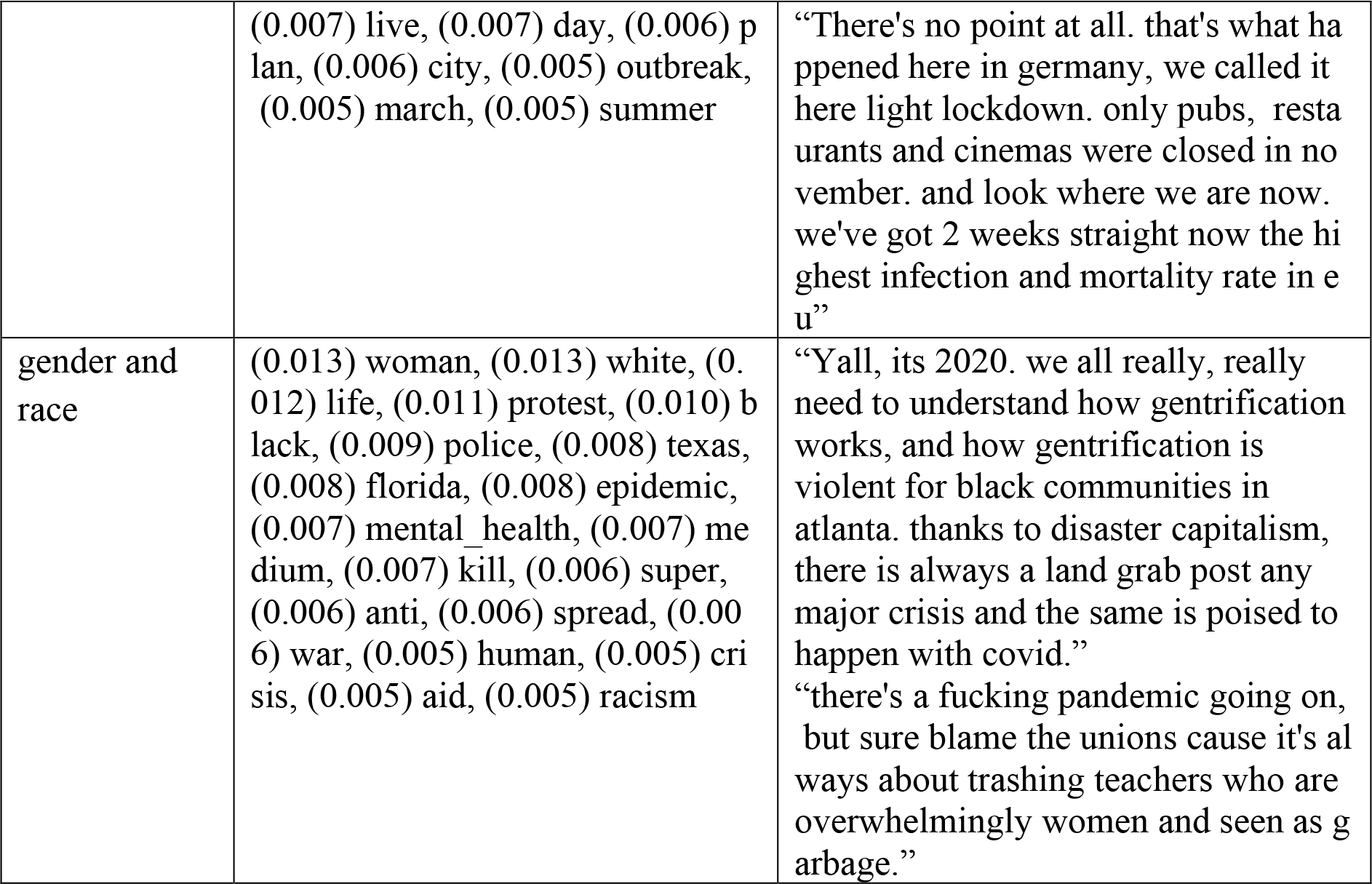
12 topics given by the LDA model and associated top-20 keywords.

**Table S4.** Topic distribution over different random samples.

|  | **Original sample (%) (n=1,742,385)** | **Sample A (%) (n=1,742,444)** | **Sample B (%) (n=3,484,880)** | **P value\*** |
| --- | --- | --- | --- | --- |
| **Topics** |  |  |  | 0.081 |
| Vaccine | 206,950 (11.88) | 205,159 (11.77) | 412,512 (11.84) |  |
| Friends and family | 228,812 (13.13) | 229,475 (13.17) | 458,420 (13.15) |  |
| Economy | 139,718 (8.02) | 138,947 (7.97) | 278,209 (7.98) |  |
| COVID-19 positive | 119,737 (6.87) | 119,589 (6.86) | 239,316 (6.87) |  |
| Medicine | 131,090 (7.52) | 130,783 (7.51) | 261,970 (7.52) |  |
| Entertainment | 95,454 (5.48) | 95,876 (5.50) | 192,091 (5.51) |  |
| Death | 103,626 (5.95) | 103,769 (5.96) | 207,392 (5.95) |  |
| Wear masks | 144,446 (8.30) | 144,854 (8.31) | 289,289 (8.30) |  |
| Children and education | 53,268 (3.06) | 53,871 (3.10) | 107,920 (3.10) |  |
| Politics | 195,601 (11.23) | 195,544 (11.22) | 391,289 (11.23) |  |
| Lockdown | 218,585 (12.55) | 218,394 (12.53) | 436,592 (12.53) |  |
| Gender and race | 105,098 (6.03) | 106,183 (6.10) | 209,880 (6.02) |  |

\*P values were calculated using $\chi^2$ test.

**Table S5.**
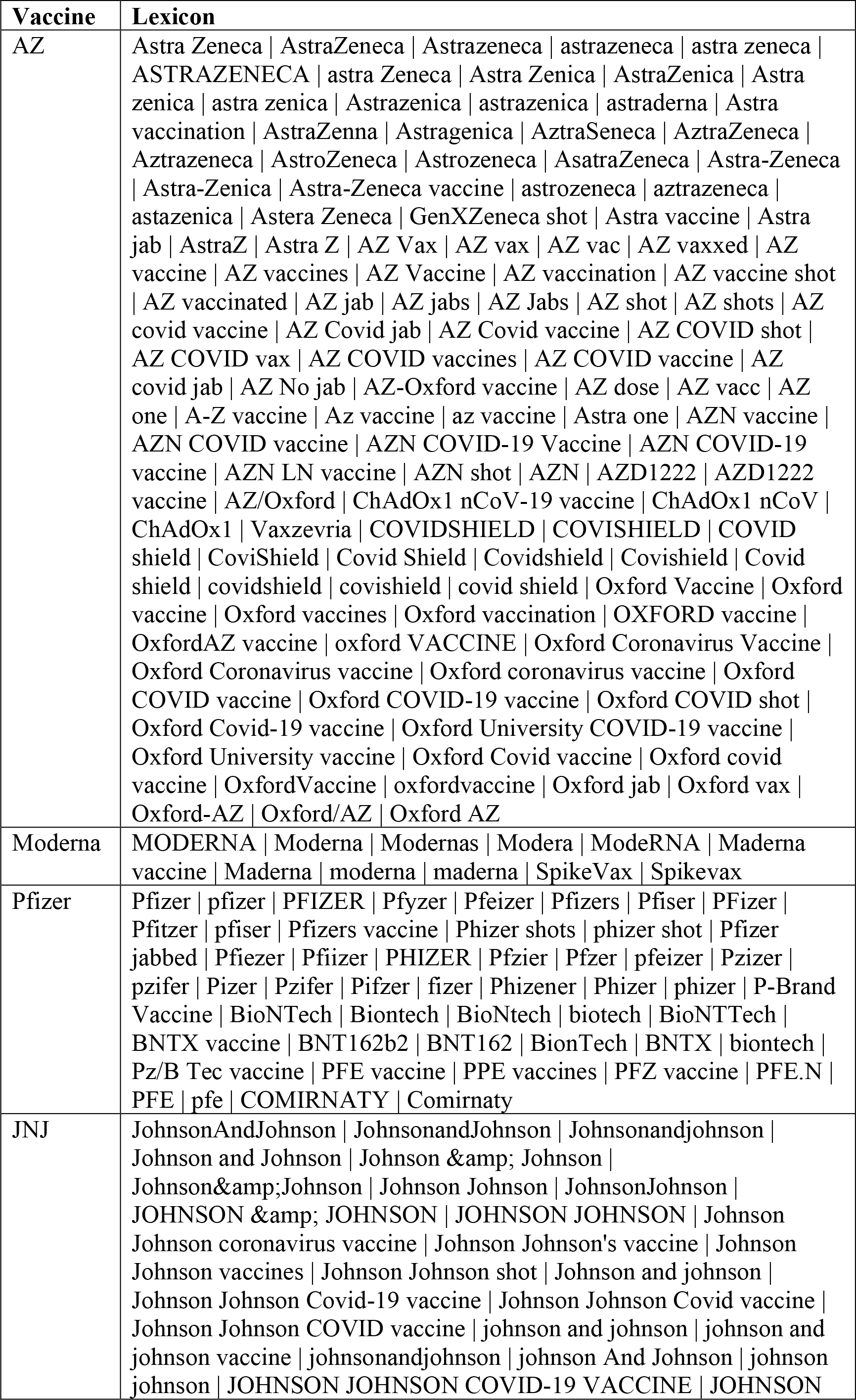
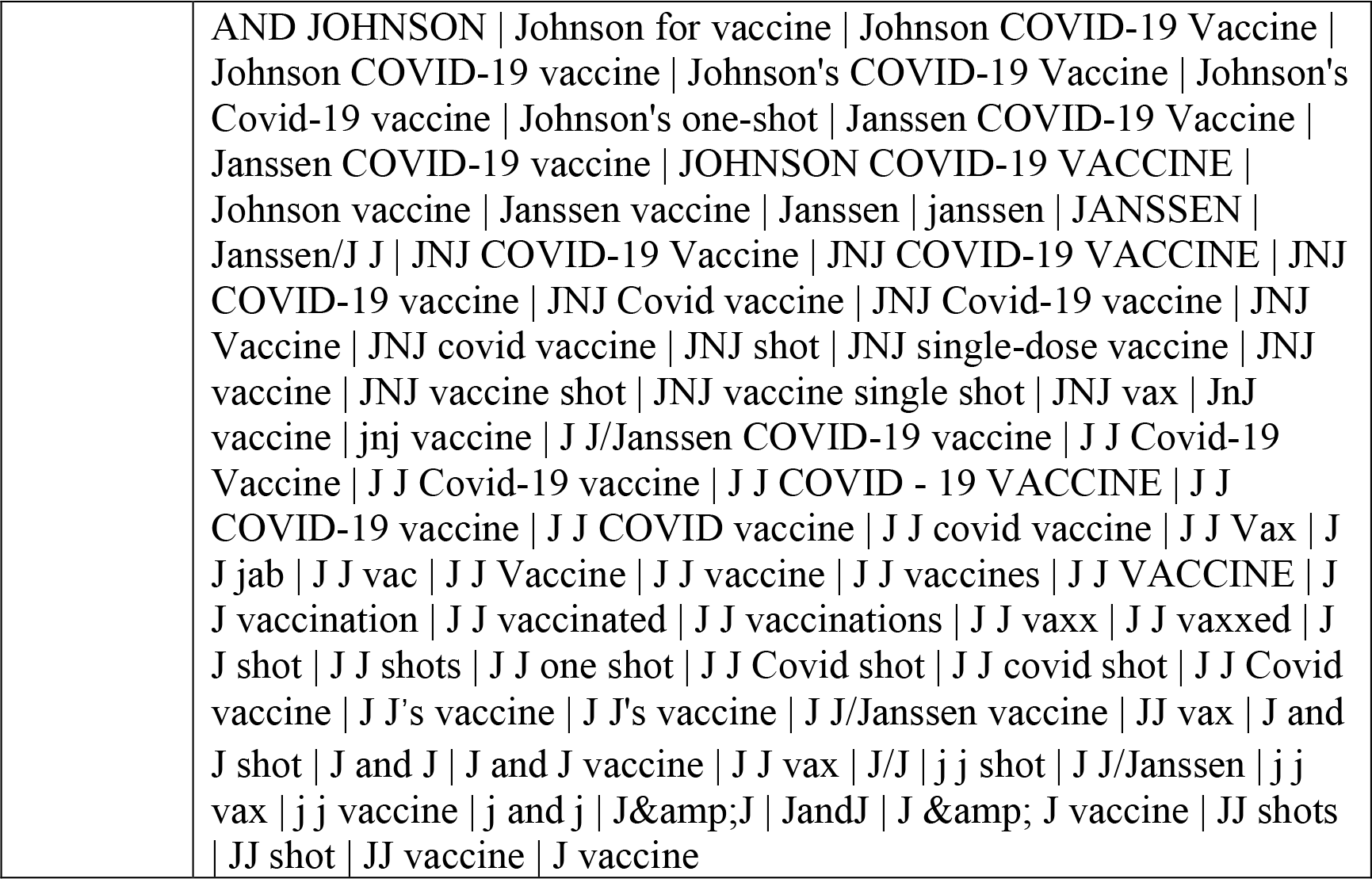
Lexicon for vaccine incorporation.

**Table S6.** Keywords for the selection of users who self-reported positive. Users who post word combinations from the “Verb” and “Noun” columns or keywords from the “Other” column on Twitter are identified as infected.

| Verb | Noun | Other |
| --- | --- | --- |
| get \| got \| have \| had \|<br>diagnose \| diagnosed \|<br>diagnose with \| diagnosed<br>with \| catch \| caught by \|<br>infected by | covid \| corona \| ncov \|<br>covid-19 \| covid19 \|<br>coronavirus \| koronavirus<br>\| sars-cov-2 \| covd \| virus \|<br>a virus \| the virus | was positive \| were<br>positive \| test positive \|<br>tested positive \| identified<br>by test \| recognized by test |

## References

1. Marmot, M. and J. Allen, COVID-19: exposing and amplifying inequalities. J Epidemiol Community Health, 2020. 74(9): p. 681–682.

2. Gibb, J.K., L.Z. DuBois, S. Williams, L. McKerracher, R.-P. Juster, and J. Fields, Sexual and gender minority health vulnerabilities during the COVID-19 health crisis. American Journal of Human Biology, 2020. 32(5): p. e23499.

3. Phillips Ii, G., D. Felt, M.M. Ruprecht, X. Wang, J. Xu, E. Pérez-Bill, R.M. Bagnarol, J. Roth, C.W. Curry, and L.B. Beach, Addressing the disproportionate impacts of the COVID-19 pandemic on sexual and gender minority populations in the United States: actions toward equity. LGBT health, 2020. 7(6): p. 279–282.

4. Beach, L.B., T.A. Elasy, and G. Gonzales, Prevalence of self-reported diabetes by sexual orientation: Results from the 2014 Behavioral Risk Factor Surveillance System. Lgbt Health, 2018. 5(2): p. 121–130.

5. O’Neill, K., Health vulnerabilities to COVID-19 among LGBT adults in California. 2020.

6. Heslin, K.C., Sexual orientation disparities in risk factors for adverse COVID-19–related outcomes, by race/ethnicity—Behavioral Risk Factor Surveillance System, United States, 2017–2019. MMWR. Morbidity and Mortality Weekly Report, 2021. 70.

7. Konnoth, C., Supporting LGBT communities in the COVID-19 pandemic. 2020). Assessing Legal Responses to COVID-19. Boston: Public Health Law Watch, U of Colorado Law Legal Studies Research Paper, 2020(20-47).

8. Anderson, R.M., C. Vegvari, J. Truscott, and B.S. Collyer, Challenges in creating herd immunity to SARS-CoV-2 infection by mass vaccination. The Lancet, 2020. 396(10263): p. 1614–1616.

9. Britton, T., F. Ball, and P. Trapman, A mathematical model reveals the influence of population heterogeneity on herd immunity to SARS-CoV-2. science, 2020. 369(6505): p. 846–849.

10. Garg, I., H. Hanif, N. Javed, R. Abbas, S. Mirza, M.A. Javaid, S. Pal, R. Shekhar, and A.B. Sheikh, COVID-19 vaccine hesitancy in the LGBTQ+ population: a systematic review. Infectious Disease Reports, 2021. 13(4): p. 872–887.

11. Carson, S.L., A. Casillas, Y. Castellon-Lopez, L.N. Mansfield, J. Barron, E. Ntekume, R. Landovitz, S.D. Vassar, K.C. Norris, and S.M. Dubinett, COVID-19 vaccine decision-making factors in racial and ethnic minority communities in Los Angeles, California. JAMA network open, 2021. 4(9): p. e2127582–e2127582.

12. Machingaidze, S. and C.S. Wiysonge, Understanding COVID-19 vaccine hesitancy. Nature medicine, 2021. 27(8): p. 1338–1339.

13. Gonzales, G., E.L. de Mola, K.A. Gavulic, T. McKay, and C. Purcell, Mental health needs among lesbian, gay, bisexual, and transgender college students during the COVID-19 pandemic. Journal of Adolescent Health, 2020. 67(5): p. 645–648.

14. Adamson, T., M. Hanley, S. Baral, C. Beyrer, S. Wallach, and S. Howell, Rapid, application-based survey to characterise the impacts of COVID-19 on LGBTQ+ communities around the world: an observational study. BMJ open, 2022. 12(4): p. e041896.

15. Lynch, K.E., J.C. Shipherd, E. Gatsby, B. Viernes, S.L. DuVall, and J.R. Blosnich, Sexual orientation-related disparities in health conditions that elevate COVID-19 severity. Annals of Epidemiology, 2022. 66: p. 5–12.

16. Rivera, A.S., M. Plank, A. Davis, M.J. Feinstein, L.K. Rusie, and L.B. Beach, Assessing widening disparities in HbA1c and systolic blood pressure retesting during the COVID-19 pandemic in an LGBTQ+-focused federally qualified health center in Chicago: a retrospective cohort study using electronic health records. BMJ Open Diabetes Research and Care, 2022. 10(6): p. e002990.

17. Wu, J., X. Liu, M. Li, W. Li, Z. Su, S. Lin, L. Garay, Z. Zhang, Y. Zhang, and Q. Zeng, Clinical text datasets for medical artificial intelligence and large language models—a systematic review. NEJM AI, 2024: p. AIra2400012.

18. Rains, S.A., Big data, computational social science, and health communication: a review and agenda for advancing theory. Health communication, 2020. 35(1): p. 26–34.

19. Jordan, S.E., S.E. Hovet, I.C.-H. Fung, H. Liang, K.-W. Fu, and Z.T.H. Tse, Using Twitter for public health surveillance from monitoring and prediction to public response. Data, 2018. 4(1): p. 6.

20. Wu, J., X. Wu, Y. Hua, S. Lin, Y. Zheng, and J. Yang. Exploring social media for early detection of depression in covid-19 patients. in Proceedings of the ACM Web Conference 2023. 2023.

21. Tsao, S.-F., H. Chen, T. Tisseverasinghe, Y. Yang, L. Li, and Z.A. Butt, What social media told us in the time of COVID-19: a scoping review. The Lancet Digital Health, 2021. 3(3): p. e175–e194.

22. Zhang, Y., H. Lyu, Y. Liu, X. Zhang, Y. Wang, and J. Luo, Monitoring depression trends on Twitter during the COVID-19 pandemic: observational study. JMIR infodemiology, 2021. 1(1): p. e26769.

23. Crocamo, C., M. Viviani, L. Famiglini, F. Bartoli, G. Pasi, and G. Carrà, Surveilling COVID-19 emotional contagion on twitter by sentiment analysis. European Psychiatry, 2021. 64(1): p. e17.

24. Sinnenberg, L., A.M. Buttenheim, K. Padrez, C. Mancheno, L. Ungar, and R.M. Merchant, Twitter as a tool for health research: a systematic review. American journal of public health, 2017. 107(1): p. e1–e8.

25. Shah, Z., D. Surian, A. Dyda, E. Coiera, K.D. Mandl, and A.G. Dunn, Automatically appraising the credibility of vaccine-related web pages shared on social media: a Twitter surveillance study. Journal of medical Internet research, 2019. 21(11): p. e14007.

26. Hua, Y., H. Jiang, S. Lin, J. Yang, J.M. Plasek, D.W. Bates, and L. Zhou, Using Twitter data to understand public perceptions of approved versus off-label use for COVID-19-related medications. Journal of the American Medical Informatics Association, 2022. 29(10): p. 1668–1678.

27. Hamamsy, T. and R. Bonneau, Twitter activity about treatments during the COVID-19 pandemic: case studies of remdesivir, hydroxychloroquine, and convalescent plasma. medRxiv, 2020: p. 2020.06. 18.20134668.

28. Chew, C. and G. Eysenbach, Pandemics in the age of Twitter: content analysis of Tweets during the 2009 H1N1 outbreak. PloS one, 2010. 5(11): p. e14118.

29. Masri, S., J. Jia, C. Li, G. Zhou, M.-C. Lee, G. Yan, and J. Wu, Use of Twitter data to improve Zika virus surveillance in the United States during the 2016 epidemic. BMC public health, 2019. 19: p. 1–14.

30. Müller, M., M. Salathé, and P.E. Kummervold, Covid-twitter-bert: A natural language processing model to analyse covid-19 content on twitter. Frontiers in artificial intelligence, 2023. 6: p. 1023281.

31. Jiang, H., Y. Hua, D. Beeferman, and D. Roy, Annotating the Tweebank corpus on named entity recognition and building NLP models for social media analysis. arXiv preprint arXiv:2201.07281, 2022.

32. Zhou, P., Z. Wang, D. Chong, Z. Guo, Y. Hua, Z. Su, Z. Teng, J. Wu, and J. Yang, Mets-cov: A dataset of medical entity and targeted sentiment on covid-19 related tweets. Advances in Neural Information Processing Systems, 2022. 35: p. 21916–21932.

33. Li, W., Y. Hua, P. Zhou, Z. Li, X. Xu, and J. Yang, Characterizing Public Sentiments and Drug Interactions during COVID-19: A Pretrained Language Model and Network Analysis of Social Media Discourse. medRxiv, 2024: p. 2024.06. 06.24308537.

34. Chen, E., K. Lerman, and E. Ferrara, Tracking social media discourse about the covid-19 pandemic: Development of a public coronavirus twitter data set. JMIR public health and surveillance, 2020. 6(2): p. e19273.

35. Mayer, K.H., J.B. Bradford, H.J. Makadon, R. Stall, H. Goldhammer, and S. Landers, Sexual and gender minority health: what we know and what needs to be done. American journal of public health, 2008. 98(6): p. 989–995.

36. Blei, D.M., A.Y. Ng, and M.I. Jordan, Latent dirichlet allocation. Journal of machine Learning research, 2003. 3(Jan): p. 993–1022.

37. Řehůřek, R. and P. Sojka, Software framework for topic modelling with large corpora. 2010.

38. Sievert, C. and K. Shirley. LDAvis: A method for visualizing and interpreting topics. in Proceedings of the workshop on interactive language learning, visualization, and interfaces. 2014.

39. Kessler, J.S., Scattertext: a browser-based tool for visualizing how corpora differ. arXiv preprint arXiv:1703.00565, 2017.

40. Chen, C., Z. Teng, and Y. Zhang. Inducing target-specific latent structures for aspect sentiment classification. in Proceedings of the 2020 conference on empirical methods in natural language processing (EMNLP). 2020.

41. Devlin, J., M.-W. Chang, K. Lee, and K. Toutanova, Bert: Pre-training of deep bidirectional transformers for language understanding. arXiv preprint arXiv:1810.04805, 2018.

42. Yang, J. and Y. Zhang, NCRF++: An open-source neural sequence labeling toolkit. In Proceedings of ACL 2018, System Demonstrations (pp. 74–79).

43. Wu, J., L. Wang, Y. Hua, M. Li, L. Zhou, D.W. Bates, and J. Yang, Trend and co-occurrence network of COVID-19 symptoms from large-scale social media data: Infoveillance study. Journal of Medical Internet Research, 2023. 25: p. e45419.

44. Li, M., Y. Hua, Y. Liao, L. Zhou, X. Li, L. Wang, and J. Yang, Tracking the impact of covid-19 and lockdown policies on public mental health using social media: Infoveillance study. Journal of Medical Internet Research, 2022. 24(10): p. e39676.

45. Thelwall, M. and S. Thelwall, Twitter during COVID-19: George Floyd opening a space to address systematic and institutionalized racism? Available at SSRN 3764867, 2021.

46. Reny, T.T. and B.J. Newman, The opinion-mobilizing effect of social protest against police violence: Evidence from the 2020 George Floyd protests. American political science review, 2021. 115(4): p. 1499–1507.

47. Baccini, L., A. Brodeur, and S. Weymouth, The COVID-19 pandemic and the 2020 US presidential election. Journal of population economics, 2021. 34: p. 739–767.

48. Sears, B., K.J. Conron, and A.R. Flores, The impact of the fall 2020 COVID-19 surge on LGBT adults in the US. 2021.

49. Tahir, A., L. Cheng, P. Sheth, and H. Liu, Improving vaccine stance detection by combining online and offline data. arXiv preprint arXiv:2208.04491, 2022.

50. Lyu, H., J. Wang, W. Wu, V. Duong, X. Zhang, T.D. Dye, and J. Luo, Social media study of public opinions on potential COVID-19 vaccines: informing dissent, disparities, and dissemination. Intelligent medicine, 2022. 2(01): p. 1–12.

51. Kuehn, B.M., Racial and Ethnic, Gender Disparities Seen in LGBT COVID-19 Vaccination Rates. JAMA, 2022. 327(10): p. 910–910.

52. McNaghten, A., COVID-19 vaccination coverage and vaccine confidence by sexual orientation and gender identity—United States, August 29–October 30, 2021. MMWR. Morbidity and Mortality Weekly Report, 2022. 71.

53. Mallory, C., B. Sears, and A. Flores, COVID-19 and LGBT Adults Ages 45 and Older in the US. 2021.

54. Teixeira da Silva, D., K. Biello, W.Y. Lin, P.K. Valente, K.H. Mayer, L. Hightow-Weidman, and J.A. Bauermeister, COVID-19 vaccine acceptance among an online sample of sexual and gender minority men and transgender women. Vaccines, 2021. 9(3): p. 204.

55. Jaiswal, J., K.D. Krause, R.J. Martino, P.A. D’Avanzo, M. Griffin, C.B. Stults, A.G. Karr, and P.N. Halkitis, SARS-CoV-2 vaccination hesitancy and behaviors in a national sample of people living with HIV. AIDS patient care and STDs, 2021. 36(1): p. 34–44.

56. Aiyegbusi, O.L., S.E. Hughes, G. Turner, S.C. Rivera, C. McMullan, J.S. Chandan, S. Haroon, G. Price, E.H. Davies, and K. Nirantharakumar, Symptoms, complications and management of long COVID: a review. Journal of the Royal Society of Medicine, 2021. 114(9): p. 428–442.

57. Han, Q., B. Zheng, L. Daines, and A. Sheikh, Long-term sequelae of COVID-19: a systematic review and meta-analysis of one-year follow-up studies on post-COVID symptoms. Pathogens, 2022. 11(2): p. 269.

58. Huang, Y., M.D. Pinto, J.L. Borelli, M. Asgari Mehrabadi, H.L. Abrahim, N. Dutt, N. Lambert, E.L. Nurmi, R. Chakraborty, and A.M. Rahmani, COVID symptoms, symptom clusters, and predictors for becoming a long-hauler looking for clarity in the haze of the pandemic. Clinical nursing research, 2022. 31(8): p. 1390–1398.

59. Zhang, H., C. Zang, Z. Xu, Y. Zhang, J. Xu, J. Bian, D. Morozyuk, D. Khullar, Y. Zhang, and A.S. Nordvig, Data-driven identification of post-acute SARS-CoV-2 infection subphenotypes. Nature Medicine, 2023. 29(1): p. 226–235.

60. Sahin, T., A. Ayyildiz, K. Gencer-Atalay, C. Akgün, H.M. Özdemir, and B. Kuran, Pain symptoms in COVID-19. American journal of physical medicine & rehabilitation, 2021. 100(4): p. 307–312.

61. Weng, L.-M., X. Su, and X.-Q. Wang, Pain symptoms in patients with coronavirus disease (COVID-19): a literature review. Journal of Pain Research, 2021: p. 147–159.

62. Wood, C.I., Z. Yu, D.-A. Sealy, I. Moss, E. Zigbuo-Wenzler, C. McFadden, D. Landi, and A.M. Brace, Mental health impacts of the COVID-19 pandemic on college students. Journal of American college health, 2024. 72(2): p. 463–468.

63. Li, Y., Q. Qin, Q. Sun, L.D. Sanford, A.N. Vgontzas, and X. Tang, Insomnia and psychological reactions during the COVID-19 outbreak in China. Journal of Clinical Sleep Medicine, 2020. 16(8): p. 1417–1418.

64. Voitsidis, P., I. Gliatas, V. Bairachtari, K. Papadopoulou, G. Papageorgiou, E. Parlapani, M. Syngelakis, V. Holeva, and I. Diakogiannis, Insomnia during the COVID-19 pandemic in a Greek population. Psychiatry research, 2020. 289: p. 113076.

65. Morin, C.M., L.-A. Vézina-Im, H. Ivers, J.-A. Micoulaud-Franchi, P. Philip, M. Lamy, and J. Savard, Prevalent, incident, and persistent insomnia in a population-based cohort tested before (2018) and during the first-wave of COVID-19 pandemic (2020). Sleep, 2022. 45(1): p. zsab258.

66. Salerno, J.P., N.D. Williams, and K.A. Gattamorta, LGBTQ populations: Psychologically vulnerable communities in the COVID-19 pandemic. Psychological trauma: Theory, research, practice, and policy, 2020. 12(S1): p. S239.

67. Zhai, Y. and X. Du, Disparities and intersectionality in social support networks: addressing social inequalities during the COVID-19 pandemic and beyond. Humanities and Social Sciences Communications, 2022. 9(1): p. 1–5.

68. Mislove, A., S. Lehmann, Y.-Y. Ahn, J.-P. Onnela, and J. Rosenquist. Understanding the demographics of Twitter users. in Proceedings of the international AAAI conference on web and social media. 2011.

69. Hua, Y., J. Wu, S. Lin, M. Li, Y. Zhang, D. Foer, S. Wang, P. Zhou, J. Yang, and L. Zhou, Streamlining social media information extraction for public health research with deep learning. Journal of the American Medical Informatics Association, 2024: p. ocae118.

